# Opposing and emotion-specific frontal alterations during facial emotion processing in generalized anxiety and depression

**DOI:** 10.1101/2022.05.18.22275191

**Authors:** Yuanshu Chen, Congcong Liu, Fei Xin, Zhou Haocen, Yulan Huang, Jinyu Wang, Jing Dai, Zhili Zou, Stefania Ferraro, Keith M Kendrick, Bo Zhou, Xiaolei Xu, Benjamin Becker

**Affiliations:** The Clinical Hospital of Chengdu Brain Science Institute, MOE Key Laboratory for NeuroInformation, University of Electronic Science and Technology of China, Chengdu, Sichuan 610054, China; Department of Psychology, Xinxiang Medical University, Henan 453003, China; School of Psychology, Shenzhen University, Shenzhen, China; Department of Psychosomatic Medicine, Sichuan Academy of Medical Sciences & Sichuan Provincial People’s Hospital, Chengdu, Sichuan, 610072, China; School of Psychology, Shandong Normal University, Jinan, 250358, China

**Keywords:** Anxiety, Depression, Generalized Anxiety Disorder, fMRI, Emotion

## Abstract

**Background:** Major depression (MDD) and generalized anxiety disorder (GAD) have become one of the leading global causes of disability and both are characterized by marked interpersonal and social impairments. However, despite a high comorbidity and overlapping social-emotional deficits it remains unclear whether MDD and GAD share a common neural basis during interpersonal processing.

**Methods:** This study combined an emotional face processing paradigm with fMRI and dimensional and categorical analyses in a sample of unmedicated MDD and GAD patients (N = 72) as well as healthy controls (N = 35).

**Results:** No group differences were found in categorical analyses. However, the dimensional analyses revealed that dorsolateral prefrontal cortex (dlPFC) reactivity to sad facial expressions was positively associated with depressive, yet negatively associated with GAD symptom load in the entire sample. On the network level depression symptom load was positively associated with functional connectivity between the bilateral amygdala and a widespread network including the anterior cingulate and insular cortex.

**Limitations:** Sex differences were not examined in the present study and some patients exhibited depression-GAD comorbidity.

**Conclusions:** Together, these findings suggest that the dlPFC - engaged in cognitive and emotional processing - exhibits symptom- and emotion-specific alteration during interpersonal processing. Dysregulated communication between amygdala and core regions of the salience network may represent MDD-specific neural dysregulations.

## Introduction

Symptomatic and epidemiological perspectives suggest that generalized anxiety disorder (GAD) and major depressive disorder (MDD) share symptoms of general affective distress and exhibit high rates of co-morbidity (70-90% lifetime comorbidity) leading to ongoing debates about common and distinct neurobiological bases (Maron and Nutt, 2017). Determining disorder-specific neural alterations in GAD and MDD can help understanding of the pathogenesis of these disorders and inform the development of more efficient interventions (MacNamara et al., 2017; Liu et al., 2021). An increasing number of studies have therefore adopted a transdiagnostic neuroimaging approach employing a direct comparison between GAD and MDD in order to determine common and disorder-specific neural alterations.

On the symptomatic level, both MDD and GAD are characterized by dysfunctional emotional and impaired cognitive processing which may in turn lead to poor social functioning (Ladouceur et al., 2005; Mathews and MacLeod, 2005; Ruscio et al., 2011). Accumulating evidence indicates that emotional dysregulations in particular represent a common core deficit in GAD and MDD, with case control studies suggesting that biased attention for negative-valence stimuli, including stimuli in social contexts such as facial expressions, may underlie the maintenance and exacerbation of negative emotional states in both disorders (Bourke et al., 2010; Foland-Ross and Gotlib, 2012; Mogg and Bradley, 2005). Neuroimaging case control studies additionally suggest altered neural processing of negatively valenced stimuli, including faces with sad or fearful expressions (Blair et al., 2008; Blom et al., 2015; Palm et al., 2011; Stuhrmann et al., 2011) in both GAD and MDD.

Functional magnetic resonance imaging (fMRI) studies suggest that intact emotional face processing relies on the visual system and a cognitive-affective circuit, including the amygdala and prefrontal regions (Fusar-Poli et al., 2009; Gur et al., 2002; Sabatinelli et al., 2011). Previous neuroimaging case control studies consistently reported increased amygdala activation in MDD patients during negative facial as well as general negative-valence emotional stimuli processing (Groenewold et al., 2013; Hamilton et al., 2012; Stuhrmann et al., 2011). Moreover, in comparison to healthy controls MDD patients exhibited altered functional communication between the amygdala and prefrontal regions during the resting state (Pannekoek et al., 2015) and the amygdala and prefrontal or insular cortex during interpersonal emotion processing (Kong et al., 2013; Xu et al., 2020). Similar with depression, intra-amygdala perturbation and abnormal connectivity of the amygdala during the resting state and emotional processing has been reported in GAD patients (Etkin et al., 2009; Li et al., 2020; Xu et al., 2021). With respect to facial emotion processing-related amygdala activation previous studies revealed inconsistent results in GAD such that both elevated and decreased amygdala activity have been reported in response to emotional faces (Blair et al., 2008; Duval et al., 2015; Fonzo et al., 2015; Monk et al., 2008).

In addition to amygdala alterations case control studies provided evidence for prefrontal cortex (PFC) dysregulations during face processing in MDD as well as GAD patients. Several studies reported PFC hypo-whereas others conversely revealed PFC hyper-activity during face processing in MDD patients compared with healthy controls (Frodl et al., 2009; Groenewold et al., 2013; Stuhrmann et al., 2011; Zhong et al., 2011). With respect to GAD increased PFC reactivity towards threatening (angry) facial expressions and increased cingulate and insular reactivity towards (non-social) threatening stimuli has been reported (Blair et al., 2008; Buff et al., 2016). However, several neuroimaging studies reported deficient frontal engagement in GAD which may suggest deficient emotion regulation in GAD (Ball et al., 2013; Xu et al., 2022), although other studies suggest that these deficits may be driven by depression rather than GAD per se (Liu et al., 2021).

The above neuroimaging studies provide initial evidence for common and dissociable neurofunctional alterations in MDD and GAD, however the conventional case control design comparing a single group of patients with healthy controls does not permit direct conclusions to be drawn about common and specific alterations and therefore dimensional and transdiagnostic neuroimaging approaches have been increasingly advocated (e.g. Etkin, 2019; Li et al., 2018; Luo et al., 2018; Parkes et al., 2020). These approaches are based on overarching conceptualizations such as the Research Domain Criteria (RDoC) that propose neurobiological dysregulations underlying mental disorders can be characterized on a dimensional continuum that cuts across traditional (categorical) diagnostic categories. Initial neuroimaging studies thus employed this transdiagnostic approach in MDD and GAD patients to determine disorder-specific lateral PFC alterations during implicit emotional regulation and emotional context-specific inhibitory control (Etkin and Schatzberg, 2011; Liu et al., 2021), dysregulations of dorsomedial PFC during social emotional working memory (Xu et al., 2022), insula dysfunctions during pain empathic processing (Xu et al., 2020) and network connectivity alterations of frontal regions during the resting state (Xu et al., 2021).

Against this background, the present transdiagnostic fMRI study aims at investigating the common and specific neurofunctional alterations in MDD and GAD during emotional face processing in treatment-naïve GAD (n = 35), MDD (n = 37) patients and healthy controls (n = 35). We employed both, a categorical approach comparing the treatment groups and a dimensional approach to examine the associations between depressive and anxiety symptom-load with neural activity and functional connectivity in the entire sample.

Given numerous studies revealed that depressive patients exhibited negative bias towards negative stimuli (Foland-Ross and Gotlib, 2012; Ho et al., 2014), we expected that the neural alterations in amygdala and PFC engaged in emotional processing would be observed during negative face processing. Based on previous findings describing neural alterations in amygdala-frontal circuits engaged in emotion regulation and social processing in both depression and pathological anxiety including GAD (Ball et al., 2013; Fonzo et al., 2015; Kim et al., 2011), we hypothesized differential neurofunctional dysregulations in these circuits would be particularly associated with marked alterations as a function of depressive symptom load in response to negative faces (see e.g. Blom et al., 2015; Satterthwaite et al., 2016).

## Methods

### Participants

A total of 72 unmedicated patients (GAD: n=35, MDD: n=37) were recruited from the Sichuan Provincial People’s Hospital and The Fourth People’s Hospital of Chengdu (Chengdu, China). Diagnoses were made according to DSM-IV criteria (Sichuan Provincial People’s Hospital) or ICD-10 (Fourth People’s Hospital of Chengdu) and to further confirm the diagnostic accuracy, an independent assessment was conducted using Mini International Neuropsychiatric Interview (M.I.N.I.) for DSM-IV (Sheehan et al., 1998). To avoid possible confounding effects of pharmacological or behavioral treatment and changes related to progressive maladaptations during the course of recurrent episodes of the disorders only patients free from current or regular pharmacological treatment with a first and recent diagnosis of GAD or MDD were included. All assessments were scheduled during the period of further diagnostic clarification (< 5 days after admission) and before any treatment was initiated. 35 well matched healthy controls (HC) were recruited via local advertisement in the community. All HC were free from current or past psychiatric disorders as validated by the M.I.N.I. interview. According to the exclusion criteria 27 subjects were excluded from the final sample for analysis (leading to a total n = 80, MDD: n = 25, GAD: n = 23, and HC: n = 32) (detailed exclusion criteria for patients and HC listed in **Supplementary materials**). The demographic data and questionnaire scores were provided in **Table 1**.

**Table 1.** Demographic information (mean, sd)

|  | **GAD**<br>(n = 23) | **MDD**<br>(n = 25) | **HC**<br>(n = 32) |  |  |  |  |
| --- | --- | --- | --- | --- | --- | --- | --- |
| Male<br>(N, %) | 13 (56.6%) | 4 (16%) | 11(34.4%) | $\chi^2_{(2)} = 8.655$ | $= .01^*$ | | |
|  | *M ± sd* | *M ± sd* | *M ± sd* | *F* | *GAD*<br>*vs. HC* | *MDD*<br>*vs. HC* | *GAD*<br>*vs. MDD* |
| Age<br>(years) | 29.39 ± 6.66 | 26.36 ± 6.93 | 27.00 ± 9.38 | F(2,79) = 0.973 | = 0.82 | = 1.00 | = 0.57 |
| Education<br>(years) | 14.95 ± 3.26 | 14.26 ± 3.28 | 14.19 ± 3.28 | F(2,79) = 0.386 | = 1.00 | = 1.00 | = 1.00 |
| **Questionnaire** |  |  |  |  |  |  |  |
| BDI-II<br>(N=78) | (n = 21)<br>21.38 ± 10.87 | (n = 25)<br>32.84 ± 9.79 | (n = 32)<br>4.97 ± 5.31 | F(2,77) = 75.66<br>*** | < .001<br>*** | < .001<br>*** | < .001<br>*** |
| PSWQ<br>(N=78) | (n = 21)<br>58.52 ± 10.54 | (n = 25)<br>63.56 ± 8.61 | (n = 32)<br>39.66 ± 9.25 | F(2,77) = 51.25<br>*** | < .001<br>*** | < .001<br>*** | = .23 |
| CTQ<br>(N=76) | (n = 20)<br>48.10 ± 11.80 | (n = 25)<br>53.08 ± 12.33 | (n = 31)<br>43.32 ± 9.58 | F(2,75) = 5.33<br>** | = .42 | = .005<br>** | = .42 |

PSWQ = Penn State Worry Questionnaire; BDI-II = Beck depression Inventory II; CTQ = Childhood Trauma Questionnaire. \*\* $p < 0.01$ ; \*\*\* $p < 0.001$ , Bonferroni-corrected.

The study was part of a larger project that aimed at determining common and separable neurofunctional alterations in MDD and GAD. The present study reported findings from an fMRI face processing paradigm which was preceded by a resting state fMRI assessment (Xu et al., 2021), a pain empathy paradigm (Xu et al., 2020), an emotional inhibition task (Liu et al., 2021) and a social emotional working memory task (Xu et al., 2022). To reduce the burden for the participants they were explicitly asked whether their current status (e.g. exhaustion, emotional state) allowed them to proceed with the experimental assessments (including functional MRI assessments and questionnaire assessments). However, some patients (and one in HC) were not able to complete the questionnaire assessments because of physical fatigue and the number of participants in the GAD group varies from 23 to 21 (BDI-II and PSWQ) and 23 to 20 (CTQ) respectively.

Written informed consent was obtained from all participants after they were informed clearly of the detailed procedures. The study was approved by the local ethics committee of the University of Electronic Science and Technology of China and all procedures were in compliance with the latest revision of the Declaration of Helsinki.

### Experimental procedure

Participants were administered the Beck Depression Inventory II (BDI-II) and Penn State Worry Questionnaire (PSWQ) to assess continuous symptom load of MDD and GAD respectively. To control for previously reported effects of early life stress on the brain (e.g. Liu et al., 2020; Teicher et al., 2016) the Childhood Trauma Questionnaire (CTQ) was administered. All subjects underwent a previously evaluated facial emotion processing paradigm during fMRI (see Domes et al., 2007; Knyazev et al., 2009; Kou et al., 2020). Participants were presented with 50 grayscale facial stimuli displaying angry, fearful, sad, neutral and happy expressions (50% females) selected from the Chinese Affective Picture System (Huang and Luo, 2004), interspersed with jitter fixations (3.5-4.4 s), and were instructed to identify the gender of each face within 2.5 seconds while passively watching these faces (i.e. implicit emotional processing) (details see **Figure 1**).

**Figure 1.** fMRI and post-fMRI experimental paradigms. (A) During fMRI, subjects were instructed to indicate the gender of the facial stimuli. (B) Post-fMRI participants underwent a memory (recognition) task for the facial stimuli and provided emotional ratings in terms of emotion category recognition and emotional intensity. Note: for the preprint version the figure displaying faces in the paradigm has been removed. The figure can be obtained via request from the corresponding authors.

Following the fMRI task subjects underwent a surprise memory task for the stimuli and provided emotion category recognition and intensity ratings. For the memory task, the presented faces were intermixed with 50 new emotion facial stimuli and participants were required to judge whether they had seen each face before (forced choice by pressing the ‘F’ - old or the ‘J’ - new). Next, the faces presented during the fMRI task were presented again and subjects were asked to choose the correct emotional category (1-neutral, 2-sad, 3-happy, 4-angry and 5-fear) and rate the intensity of the emotion for each stimulus from 1- very weak to 9 - very strong (details **see Figure 1**).

### MRI data acquisition and analyses

Images were acquired using a 3.0 Tesla GE Discovery MR750 system (General Electric Medical System, Milwaukee, WI, USA) and processed using SPM12 toolbox (Friston et al., 1994) (details see **Supplementary Materials**).

On the first-level a standard general linear model (GLM) was employed to the preprocessed functional images to initially assess specific effects of task parameters on blood oxygenation level-dependent responses. The regressors of the five experimental conditions (neutral, happy, sad, angry and fearful faces) were modelled using an event-related design matrix and convolved with a standard hemodynamic response function (HRF). A temporal high-pass filter (1/128 Hz) was used. The six head motion parameters generated during the realignment step were included in the GLM as nuisance covariates. For each participant, specific contrast images for each type of facial expression were generated separately and subjected to the group level analysis.

For the second level analysis, firstly a categorical approach including the diagnostic groups was applied (details see **Supplementary Materials**). To explore whether dimensional models of depression and anxiety may explain face emotional processing alterations we next examined associations between levels of anxiety or depression and neural activation using non-parametric simple regression analyses as implemented in SnPM 13. Non-parametric tests were employed due to the violation of normal distribution (Shapiro-Wilk for BDI scores, *p* < 0.05; for PSWQ scores, *p* = 0.24). Because two patients in the GAD group did not complete the questionnaires, mean BDI / PSWQ scores for the GAD group were used to complete their data. To determine dimensional associations between depression and GAD symptom load with brain indices, whole brain voxel-wise regression analyses were conducted for BDI or PSWQ scores, with gender, PSWQ × BDI interaction score as covariates of no interest. To threshold results, a significance threshold of *p* < 0.05 (FWE) corrected at cluster level and a minimum cluster size (k) of ten voxels were set for multiple comparisons.

The physiological interaction between two brain regions can vary in different psychological contexts (Jalbrzikowski et al., 2017; Ravindranath et al., 2020) and this measure can be employed to examine alterations in the communication between brain regions in psychiatric disorders. The general Psychophysiological Interaction analysis (gPPI; McLaren et al., 2012) implements a task-dependent functional connectivity analysis based on computing correlations between the BOLD response time course of a target brain region and that of a seed region under the given psychological context, and estimates the task-dependent functional connectivity for each experimental condition using corresponding regression coefficients. We employed this approach in the present study to further examine network-level alterations in the disorders. Specifically, regions from the whole brain BOLD level activity results were defined as 6-mm sphere seed regions centered on the identified peaks in MNI space. Additionally, given previous results on altered amygdala network level integration in depression and anxiety disorders (Kolesar et al., 2019; Müller et al., 2017; Xu et al., 2020; 2021) the bilateral amygdala was defined as a seed region. For the selected pairwise seed-target pathways we conducted one-way ANOVAs for each face emotion category to evaluate the main effect of group and the interaction effect on the strength of functional connectivity. Moreover, regression analysis for the associations between the symptom dimensions and the functional connectivity was conducted and the rate of type 1 errors was corrected with a threshold of *p*_FWE_ < 0.05 (cluster level) on the whole brain level. To further disentangle the significant effects, parameter estimates were extracted for each subject using MarsBaR.

## Results

### Demographic data

Participant demographics and dimensional symptom load were presented in **Table 1**. Both patient group and healthy controls did not differ with respect to age and education level (*p*s > 0.05), but gender distribution was not matched between each group (*p* (**χ**^2^) = 0.01) with a higher proportion of females in the MDD group compared to the other two groups. One-way ANOVAs for symptom load of depression and anxiety revealed a significant main effect of group (BDI-II, F(2,77) = 75.66, *p* < 0.001; PSWQ, F(2,77) = 51.25, *p* < 0.001). Post hoc Bonferroni-corrected comparison indicated that patient group had both higher BDI and PSWQ scores than HC (*p*s < 0.001), and that participants with a primary diagnosis of MDD had higher BDI scores compared with the GAD group (*p* < 0.001), however for anxiety symptom load, there was no significant difference between the two patient groups (*p* = 0.194). For the dimensional neuroimaging analysis we additionally assessed the variance inflation factor (VIF = 2.21) to assess collinearity between anxiety and depression symptom load and results showed no potential collinearity problem (O’brien, 2007; Xu et al., 2020).

Additionally, analysis for the early life stress exposure of three groups revealed a significant main effect of group (CTQ, F(2,75) = 5.33, *p* = 0.007) with post-hoc analysis showing the CTQ scores only differed between the MDD and HC (*p* = 0.005), but not for GAD relative to HC (*p* = 42) or GAD relative to MDD patients (*p* = 0.42). Given significant group differences in these variables we repeated the analyses using gender and CTQ as covariate and results remained stable (see **Supplements**).

### Behavioral results

#### Emotional face processing

With respect to gender decision accuracy a mixed ANOVA with valence (neutral, sad, happy, angry and fearful faces) as within-subject factor and group (GAD, MDD, HC) as between-subject factor revealed no significant main effect of group (F(2,77) = 0.60, *p* = 0.55, *η*_*p*_ ^*2*^= 0.02) and valence × group interaction effect (F(8,77) = 0.93, *p* = 0.49, *η*_*p*_^*2*^ = 0.02), but a significant main effect of valence (F(4,77) = 14.34, *p* < 0.001, *η*_*p*_^*2*^ = 0.16). Post hoc Bonferroni-corrected comparison between different emotional faces showed that all participants had lower recognition accuracy for negative faces as compared to the happy (*p*s < 0.001) and neutral faces (*p*s < 0.01). A corresponding ANOVA for reaction time data also revealed a significant main effect of valence (F(4,77) = 10.35, *p* < 0.001, *η*_*p*_^*2*^ = 0.12) with longer reaction times for negative faces compared to the neutral faces (*p*s < 0.05) across all groups. Moreover, results suggested the longest RT for sad faces and only for sad faces there was a significant difference in RT compared to happy faces (sad > happy, *p* = 0.015). The main effect of group (F(2,77) = 1.90, *p* = 0.16, *η*_*p*_^*2*^ = 0.05) and the interaction effects (F(8,77) = 0.31, *p=* 0.96, *η*_*p*_^*2*^ = 0.01) were not significant (see **Figure 2**). Summarizing, both ACC and RT results suggest that all subjects attentively processed the facial stimuli and that the implicit emotion paradigm induced a modulation on the behavioral level such that negative emotion faces induced lower ACC and higher RT across all participants.

**Figure 2.**
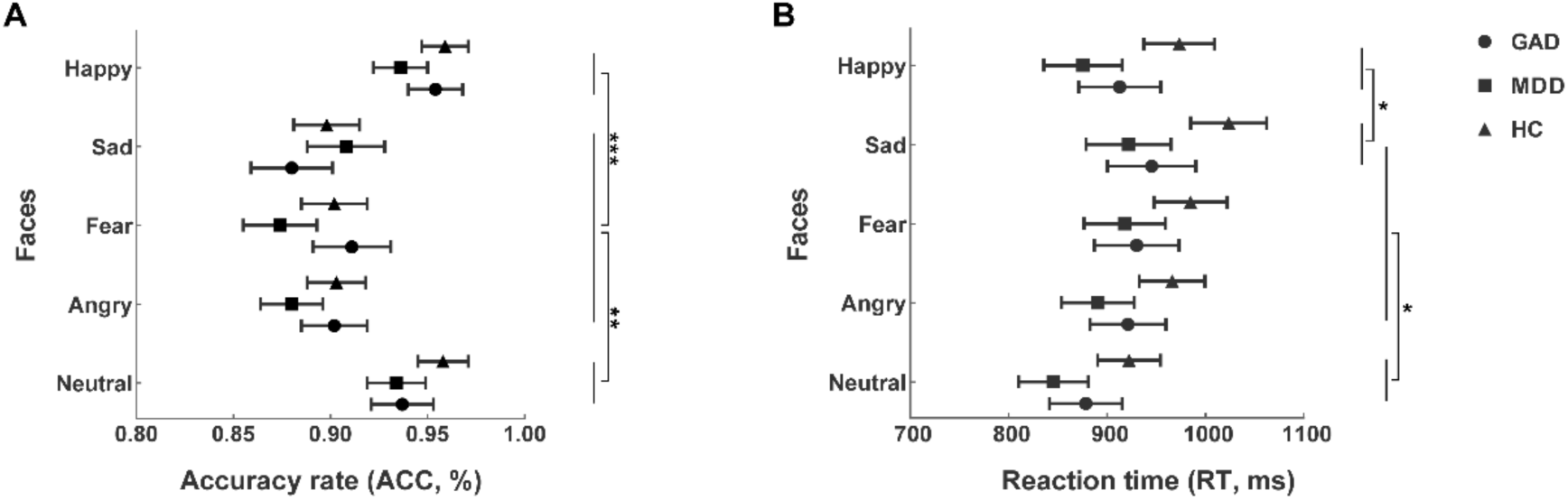
The accuracy rate (A) and reaction time (B) during the implicit face processing task. All participants (A) recognized the gender of negative faces less accurately than happy and neutral faces with (B) longer reaction time for the negative facial expressions than neutral faces. ^*\**^*p* < 0.05,^*\*\**^*p* < 0.01, ^*\*\*\**^*p* < 0.001.

To explore whether symptom load was associated with variations in the individual behavioral responses after controlling the gender, partial correlation analyses were performed which revealed no significant associations between BDI / PSWQ scores and emotion-specific accuracy (*p*s > 0.20). No significant correlations between the RT for each emotion and personal symptom load were observed (*p*s > 0.12).

### Emotional memory, emotion recognition and emotion intensity rating

Analysis of post-fMRI memory accuracy rates revealed a significant main effect of emotion condition (F(4,308) = 15.26, *p* < 0.001, *η*_*p*_^*2*^ = 0.17), with sad and fearful faces being recognized better than neutral faces (*p*s < 0.001), and the recognition of fearful faces being better than happy and angry faces (*p*s < 0.001), while there was no significant difference in recognition ACC for fearful and sad faces (*p* = 0.13). No significant main effect of group (F(2,77) = 1.07, *p* = 0.35, *η*_*p*_^*2*^ = 0.03) and interaction effects were observed (F(8,308) = 0.68, *p* = 0.71, *η*_*p*_^*2*^ = 0.02). Analysis of reaction times did not reveal any significant effects (*p*s > 0.06).

With respect to the emotional ratings (one subject failed to complete this task due to technical problems leading to N = 79), the analysis of recognition accuracy showed a significant main effect of emotion condition (F(4,304) = 20.46, *p* < 0.001, *η*_*p*_^*2*^ = 0.21) with all subjects recognizing fearful faces less accurately than other face conditions (*p*s < 0.001). The main effect of group (F(2,76) = 2.51, *p* = 0.088, *η*_*p*_^*2*^ = 0.06) and interaction effect (F(8,304) = 1.02, *p* = 0.42, *η*_*p*_^*2*^ = 0.03) were not significant. Analysis of RT revealed a significant main effect of emotion condition (F(4,304) = 53.97, *p* < 0.001, *η*_*p*_^*2*^ = 0.42) only. Consistently, all participants spent considerably longer time recognizing fearful faces compared with other types of facial expressions (*p*s < 0.001), with neutral faces being recognized quickest (*p*s < 0.001). We did not observe other significant effects of group or interactions (*p*s > 0.08). Analysis of emotion intensity ratings revealed a significant main effect of emotion condition (F(4,304) = 79.64, *p* < 0.001, *η*_*p*_^*2*^ = 0.51) with post hoc Bonferroni-corrected comparison showing that neutral and happy faces were rated less intense than negative faces (*p*s < 0.001), but no significant difference was observed in intensity ratings for sad and fearful faces (*p* = 0.98). Results did not show significant main group and interaction effects (*p* > 0.11) (see **Figure 3**).

**Figure 3.**
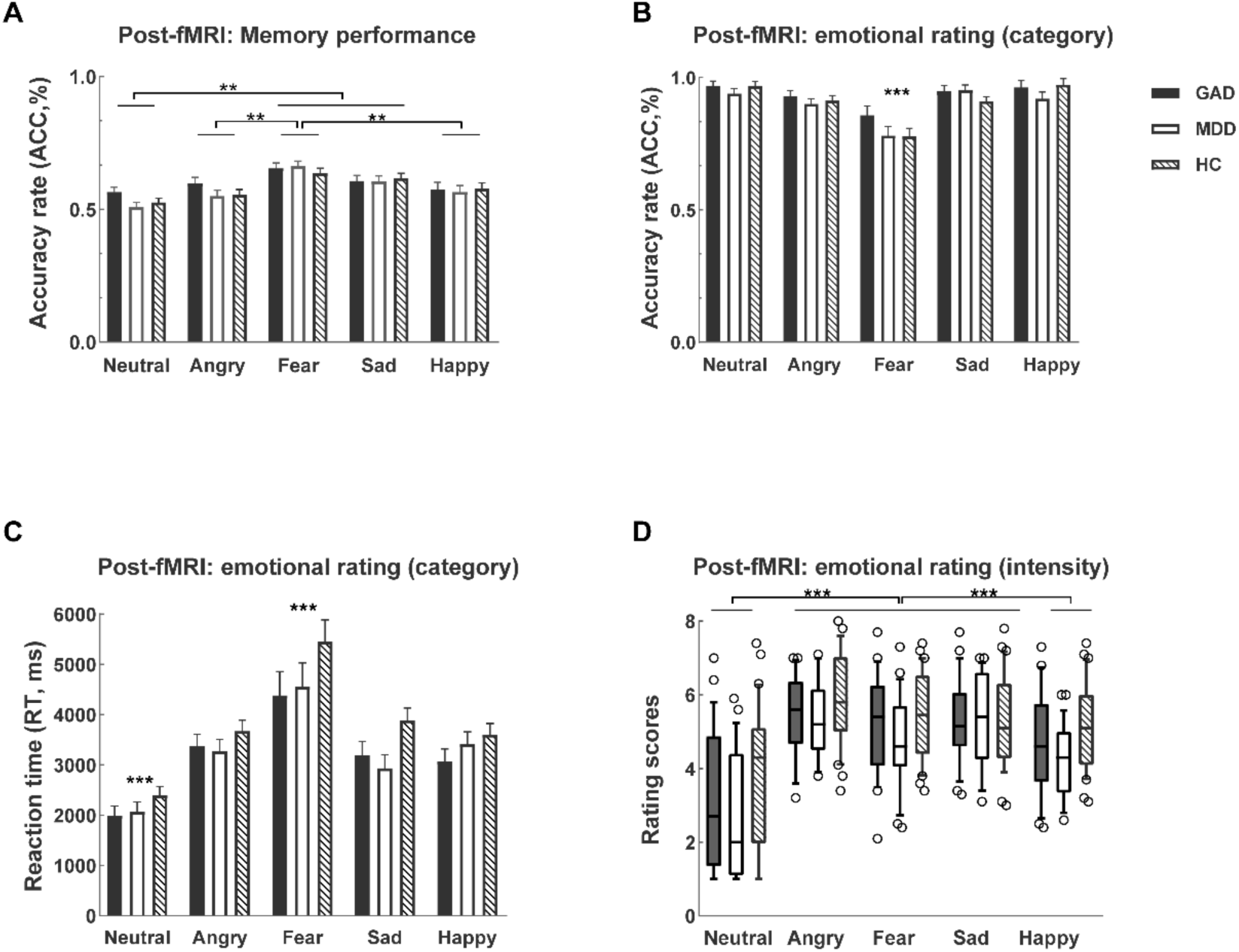
Behavioral results of the post-fMRI (A) memory and (B-D) emotional rating task. (A) Analysis for memory task performance revealed highest ACC for recognizing the fearful faces and both fearful and sad faces were memorized better than neutral faces. For the recognition task, participants (B) recognized the category of fearful faces worst (C) with longest reaction time for fearful faces, and (D) all negative faces were perceived more intense than the neural and happy faces. Histograms show mean ± SEM ACC or RT for each face condition. Error bars show standard errors. ^*\**^*p* < 0.05, ^*\*\**^*p* < 0.01, ^*\*\*\**^*p* < 0.001.

### fMRI results

#### Categorical approach: BOLD activity – group level ANOVA results

One-way ANOVA models on the whole-brain level did not find significant group differences in response to different facial expressions (*p*_FWE_ > 0.05). Additionally, amygdala-focused ROI analysis revealed no significant differences between the patient groups and healthy controls (bilateral amygdala, *p*_FWE_ > 0.05, SVC).

### Dimensional approach: BOLD activity – non - parametric regression analysis

Whole brain regression analyses revealed a significant positive association between depressive symptom load and brain activation in the left dorsolateral prefrontal cortex (dlPFC, FWE-*p*_cluster_ = 0.036, k = 105, MNI peak coordinate: -27 / 57 / 27) during processing of sad faces. In contrast, levels of GAD symptoms were found to be negatively associated with left dlPFC (FWE-*p*_cluster_ = 0.025, k = 131, MNI peak coordinate: -33 / 54 /21) activation to sad faces (see **Figure 4**). No other associations with primary diagnosis reached our threshold for significance at the whole brain level in response to other emotional faces.

**Figure 4.**
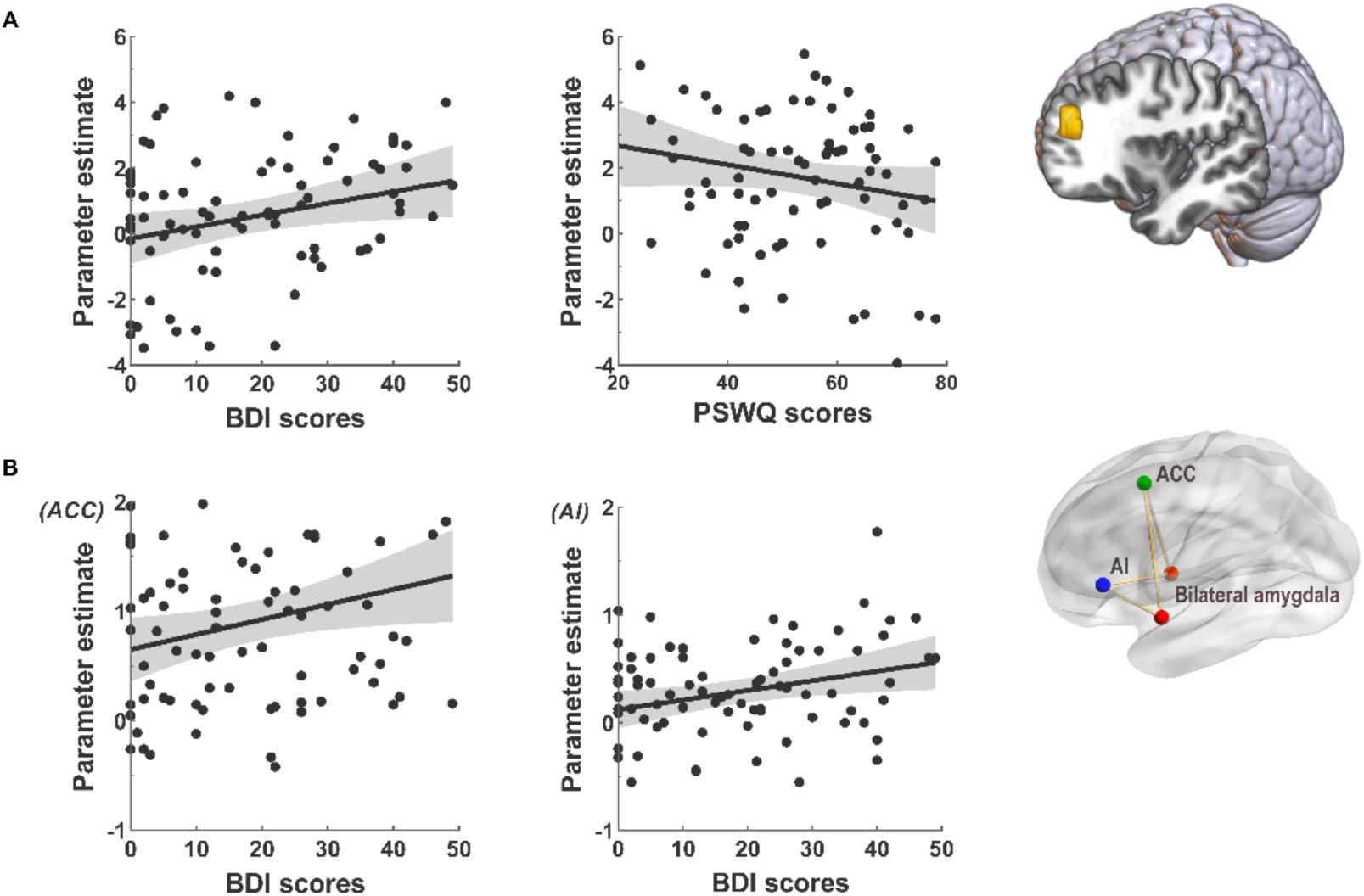
(A) Results from the whole-brain regression analysis. Neural responses in the left dlPFC to sad faces were found to be positively associated with depressive symptom load (r = 0.49, *p* < 0.001) and significantly negatively associated with anxiety symptom load (r = -0.46, *p* < 0.001). (B) Functional connectivity between bilateral amygdala and anterior cingulate cortex (ACC, r = 0.42, *p* < 0.001) and anterior insula (AI, r = 0.39, *p* < 0.001) was significantly associated with depressive symptom load in the entire sample.

### Functional connectivity analyses

For the whole brain functional connectivity analysis, both one-way ANOVA and regression models did not reveal any significant between group differences or symptom associations with respect to functional connectivity of the dlPFC during processing of sad faces (*p*_FWE_ > 0.05). Based on our priori regional hypothesis a regression model with the bilateral amygdala as seed was performed and revealed that depression level was positively associated with functional connectivity between the bilateral amygdala and anterior cingulate cortex (ACC,FWE-*p*_cluster_ = 0.031, k = 159, MNI peak coordinate: 9 / 12 / 45), and middle frontal cortex (FWE-*p*_cluster_ = 0.049, k = 112, MNI peak coordinate: -30 / 36 / 18), including the left anterior insula (AI, -27 / 30 / 3) for sad faces (see **Table 2, Figure 4**). No other significant linear associations were observed between symptoms of anxiety and depression and functional connectivity of the bilateral amygdala for other emotion conditions at the same threshold.

**Table 2.** Whole brain functional connectivity showing positive association with depression level using the bilateral amygdala seed in response to sad faces

| Brain region | Cluster k | Peak-t value | x | y | z |
| --- | --- | --- | --- | --- | --- |
| BDI: sad faces |  |  |  |  |  |
| *Positive correlation* |  |  |  |  |  |
| R. Anterior Cingulate Cortex | 159 | 4.38 | 9 | 12 | 45 |
| R. Corpus Callosum |  | 4.10 | 9 | 24 | 18 |
| R. Middle Cingulate Cortex |  | 3.89 | 15 | 24 | 30 |
| L. Middle Frontal Cortex | 112 | 4.18 | -30 | 36 | 18 |
| L. Anterior insula |  | 3.95 | -27 | 30 | 3 |
| L. Middle Frontal Cortex |  | 3.51 | -39 | 33 | 21 |

$p_{FWE} < 0.05$ , voxels $> 10$ . L indicates left; R indicates right.

## Discussion

The present study aimed to investigate common and specific neural biomarkers in GAD and MDD by employing categorical and dimensional analyses in a sample of treatment-naïve GAD and MDD patients and healthy controls. Our findings revealed dlPFC activity in response to sad facial expressions was positively associated with the level of depression symptoms, yet negatively correlated with GAD symptom level. Moreover, depressive symptom load was also positively associated with increased functional connectivity of the bilateral amygdala with the anterior cingulate cortex and anterior insula during sad face processing. In contrast, a conventional categorical analysis approach did not reveal significant differences between the diagnostic groups. The present study thus suggests that levels of GAD and MDD may be dissociable with respect to neural reactivity towards sad faces as reflected by opposite associations between dlPFC reactivity with GAD and MDD symptom load as well as MDD-specific correlation with amygdala-cingulate/insula communication.

Consistent with previous evidence suggesting that MDD patients exhibit increased activity in the dlPFC during emotional processing (Etkin and Schatzberg, 2011; Foland-Ross and Gotlib, 2012; Stuhrmann et al., 2011; however, see also Liu et al., 2021; Xu et al., 2022) the present study observed that dlPFC activation was positively associated with depressive symptoms in response to sad faces. The dlPFC represents a key region for emotion and cognitive regulation (Koenigs and Grafman, 2009; Peng et al., 2016; Zhong et al., 2016) and aberrant dlPFC neural activation in MDD was reported during working memory and cognitive control (Wagner et al., 2006; Walter et al., 2007). Overarching models and accumulating studies suggest that dysfunctional dlPFC activity and connectivity may reflect an impaired ability for top-down regulation of negative affect (Liu et al., 2021; Zimmermann et al., 2017) and positive association between levels of depression and activity in this region may signal an increased need for emotion regulation in response to sad faces. The dlPFC is a rather homogenous region and has also been associated with general attention and salience processing (Eysenck and Derakshan, 2011; Katsuki and Constantinidis, 2012). Previous studies reported an emotion-specific bias towards sad facial expressions in depression that was neurally underpinned by dlPFC alterations (e.g. Auerbach et al., 2015; Dalili et al., 2015). Within this context the present findings may reflect that biased attention for sad faces increases as a function of depressive symptom load.

On the other hand, the negative association between dlPFC activation in response to sad faces and GAD symptoms suggests a potential role of this region in the symptoms of GAD (e.g., Liu et al., 2021). The dlPFC plays an important role in cognitive control and the top-down regulation of negative emotion by means of different regulation strategies such as reappraisal, suppression and detachment (Campbell-Sills et al., 2011; Meluken et al., 2019). Together with other regions including the rostral ACC, medial PFC and hippocampus, the dlPFC has been implicated in modulating fear reactivity (Duval et al., 2015). With respect to GAD-specific neurofunctional emotional dysregulations, in line with studies showing an attenuated BOLD signal in PFC during facial affect processing (Palm et al., 2011) and emotion regulation in GAD (Ball et al., 2013), deficient recruitment of this region may thus reflect deficient dlPFC regulatory engagement in GAD during top-down emotion processing probably due to interference of worry and rumination.

Given patients suffering from anxiety and depressive disorders often show abnormal PFC functioning, prefrontal functions have been used as a treatment target in both GAD and MDD (Quidé et al., 2012). In consideration of non-invasive techniques, repetitive transcranial magnetic stimulation (rTMS) and transcranial direct current stimulation (tDCS) studies have demonstrated that stimulating the dlPFC can produce marked improvement in treatment-resistant depression (Downar and Daskalakis, 2013; Kekic et al., 2016 for a review). Similarly, stimulating the dlPFC using rTMS and tDCS, patients with GAD showed a clinically significant decrease in anxiety symptoms (Dilkov et al., 2017; Sagliano et al., 2019), suggesting a promising therapeutic potential in GAD. Interestingly, a recent rTMS study has reported that stimulation of different dlPFC locations reduce either dysphoric or anxiosomatic symptoms of MDD in treatment-resistant patients, providing some support for our findings of differential activations and locations in responses to sad faces in MDD and GAD patients (Siddiqi et al., 2020). The dlPFC may therefore be considered as a potential target for new non-invasive treatments although possibly different locations within this region may selectively influence mood and anxiety-related symptoms.

On the connectivity level, depressive symptoms were positively associated with the functional connectivity strengths between the bilateral amygdala and regions including the insula and ACC in response to sad facial expressions. As a central region in the limbic emotion-processing circuit, the amygdala is involved in facial emotion recognition, encoding and retrieval of emotional memories particularly for negative information (Foland-Ross and Gotlib, 2012; Groenewold et al., 2013; Mihov et al., 2013). Numerous studies have consistently reported increased amygdala activity in response to sad faces in MDD (Blom et al., 2015; Groenwold et al., 2013; Ho et al., 2014; Stuhrmann et al., 2011) and aberrant network connectivity of amygdala with ACC during implicit regulation of emotion processing (Etkin and Schatzberg, 2011) and with dorsal AI in MDD during pain empathy (Xu et al., 2020). The anterior insula, a core hub of the salience network (Chong et al., 2017; Koban and Pourtois, 2014), plays a key role in aversive emotional processing (Menon, 2015; Uddin, 2015; Uddin et al., 2017) while the ACC supports the integration of emotional and cognitive information (Killgore and Yurgelun-Todd, 2004; Yamasaki et al., 2002). A meta-analytic study found that MDD patients exhibited widespread dysfunction in the networks related to emotion or salience processing, internally or externally oriented attention, and goal-directed regulation functions (Kaiser et al., 2015). Within this context our findings may reflect a higher salience and preferential processing of sad facial expressions with increasing levels of depression (also see e.g. Bourke et al., 2010). Together the positive associations between amygdala-insula\ACC communication and depressive symptom during sad facial processing may indicate that MDD patients showed overall disturbed neural function involved in salience processing which consistent with previous study demonstrating that amygdala-insula and amygdala-ACC possibly contribute to the negative bias in depression.

The observed associations for both, MDD and GAD symptom load were specifically observed for sad but not other emotional face expressions. Different lines of research suggest a negative response and attention bias towards sadness-related stimuli in MDD (see Bourke et al., 2010) which might persist even after the recovery from depression (Stuhrmann et al., 2011). For instance, MDD patients tend to interpret ambiguous and neutral faces as sad and show enhanced vigilance and attention towards sad expressions (Foland-Ross and Gotlib, 2012; Gotlib et al., 2004). On the neural level, depressed patients exhibited altered activation in regions involved in emotion generation and regulation towards sad faces (Blom et al., 2015; Dichter et al., 2009). Similar with MDD adults anxiety particularly GAD patients also showed attention bias to negative stimuli including threatening faces (e.g., angry or fearful) (Blair et al., 2008; Maron and Nutt, 2017; Sylvester et al., 2016).

Finally, although we found neural alterations associated with depression or GAD symptoms we did not observe significantly reduced general or emotion-specific recognition accuracy (or slower response to emotional facial expressions) in response to emotional faces in patients compared with healthy controls. This was consistent with previous studies and could be explained by the comparable simple task that was used in the present study (Bourke et al., 2010; MacNamara et al., 2017; Palm et al., 2011).

There are some limitations in our current study. Firstly, we applied a relatively restrictive enrollment criteria in order to avoid confounding effects of treatment or progressive dysregulations during the disorder course leading to a moderate sample size. Secondly, given that previous studies have reported sex-dependent differences in depression and anxiety (Christiansen, 2015; Yao et al., 2014), sex difference should be examined in future studies with larger samples. Thirdly, though the primary diagnosis of MDD and GAD was made according to DSM-IV criteria or ICD-10, a few patients (MDD = 3 in the GAD group, GAD = 7 in the MDD group) exhibited secondary MDD or GAD comorbidity according to the M.I.N.I. interview.

## Conclusion

In the present sample of treatment-naïve GAD, MDD and healthy controls levels of MDD and GAD showed emotion-specific associations with dlPFC activation in response to sad faces. While higher levels of MDD associated with higher dlPFC activity higher levels of GAD associated with lower dlPFC activity which might reflect that the salience of sad facial expressions increases as a function of depressive symptoms, while increasing GAD levels may attenuate an implicit regulation of the emotional response towards sad facial expressions. On the network level depression symptom-specific associations with increased amygdala-insula and amygdala-ACC connectivity were observed which may reflect an increased salience of sad facial expressions. Together our results indicate sad-specific and neurofunctional separable alterations during processing of emotional faces in GAD and MDD.

## Data Availability

All data produced in the present study are available upon reasonable request to the authors

## Author contributions

Authors XX, KK and BB designed this study. YH, JD, ZZ, BZ, and JW performed the clinical assessments. YC, XX, FX, HZ and CL acquired data. Authors YC and BB conducted the statistical analysis. Author YC prepared the manuscript, and authors YC, SF, XX, BB revised the manuscript into its finalized form. All authors commented on and gave final approval to the final version of the manuscript.

## Acknowledgments

The authors would like to thank the laboratory members and colleagues for their valuable help and we also wants to appreciate all the participants in the present study.

