## Supplementary for "Opposing and emotion-specific frontal alterations during facial emotion processing in generalized anxiety and depression"

**Exclusion criteria**

Exclusion criteria applied to all participants (including healthy controls) were: (1) current or history of the following axis I disorders according to DSM criteria: bipolar, psychotic, substance use, or posttraumatic stress disorders, feeding and eating disorders, and mania, (2) current or history of medical or neurological disorders, (3) acute (within six weeks before the assessments) or chronic use of medication, (4) acute suicidal ideation, (5) contraindications for MRI assessment, (6) left handedness, and (7) excessive head motion during fMRI scanning (more than 3 mm) (see **Figure S1**).

**Behavioral data analysis**

Data from the face paradigm in terms of accuracy rate (ACC) and reaction time (RT) were examined by means of mixed-ANOVAs with group as between-subject factor and emotion category as within-subject factor. Analyses were employed in SPSS 22 (Armonk, NY: IBM Corp.). All reported p values were two tailed, and $p < 0.05$ was considered as significant.

**MRI data acquisition and preprocessing**

Images were acquired via a 3 Tesla GE MR750 system (General Electric Medical System, Milwaukee, WI, USA). The functional MRI time series were acquired using a $T2^*$-weighted Echo Planar Imaging (EPI) sequence (repetition time = 2000 ms, echo time = 30 ms, number of slices = 39, field of view = 240 × 240 mm, flip angle = 90°, image matrix = 64 × 64, thickness / gap = 3.4 / 0.6mm). Additionally, to exclude subjects with apparent brain pathologies and improve normalization accuracy during preprocessing, T1-weighted high-resolution anatomical images were acquired with a 3-dimentional spoiled gradient echo pulse sequence (repetition time = 6 ms, echo time = minimum, number of slices = 156, field of view = 256 × 256 mm, flip angle = 9°, acquisition matrix = 256 × 256, thickness = 1 mm).

Functional images were preprocessed and analyzed with SPM12 software package (Statistical Parametric Mapping; Welcome Department of Cognitive Neurology, London; http://www.fil.ion.ucl.ac.uk/spm/spm12) (Friston et al., 1994). For each run, the first six volumes of functional neuroimaging time-series were discarded to achieve magnet-steady images. Images were corrected for slice timing and head movement and normalized into the standard Montreal Neurological Institute (MNI) template space resampled at 3 × 3 × 3 mm voxel size and spatially smoothed using an 8-mm full width at half maximum (FWHM) Gaussian kernel. An examination of between-group differences in head motion as assed by mean framewise displacement revealed no significant differences ($F(2, 79) = 0.18$, $p = 0.84$).

**fMRI data analysis – categorical analysis**

On the second-level, one-way ANOVAs for each facial category were used to examine the main effects of face conditions, group differences between patients and healthy controls in processing different faces and the interactions. For the whole brain analyses, a significance threshold of $p < 0.05$ family-wise error (FWE) peak-level correction and cluster extent k ⩾ 10 was employed to account for multiple comparisons. Based on our regional-specific hypothesis additional analyses focused on the amygdala as a prior regions of interest (ROI, defined as bilateral amygdala mask according to the Automated Anatomic Labeling atlas). Within the a priori ROI results were corrected for multiple comparisons using $p < 0.05$ FWE corrected at the peak level using small volume correction (SVC).

**fMRI results**

**Non - parametric regression analysis**

Given the observed significant group difference in CTQ scores, we also tried including CTQ scores as covariate in the regression model and results remained stable that during processing of sad faces, brain activation in the left dorsal lateral prefrontal cortex (dlPFC, FWE-$p_{cluster} = 0.048$, k = 93, MNI peak coordinate: -27 / 57 / 27) was significantly positively correlated with depressive symptoms, in contrast, negatively associated with GAD level (FWE-$p_{cluster} = 0.043$, k = 98, MNI peak coordinate: -27 / 57 / 27). No other associations with primary diagnosis reached our threshold for significance at the whole brain level in response to other emotional faces.

**Figure S1. CONSORT Flow chart**

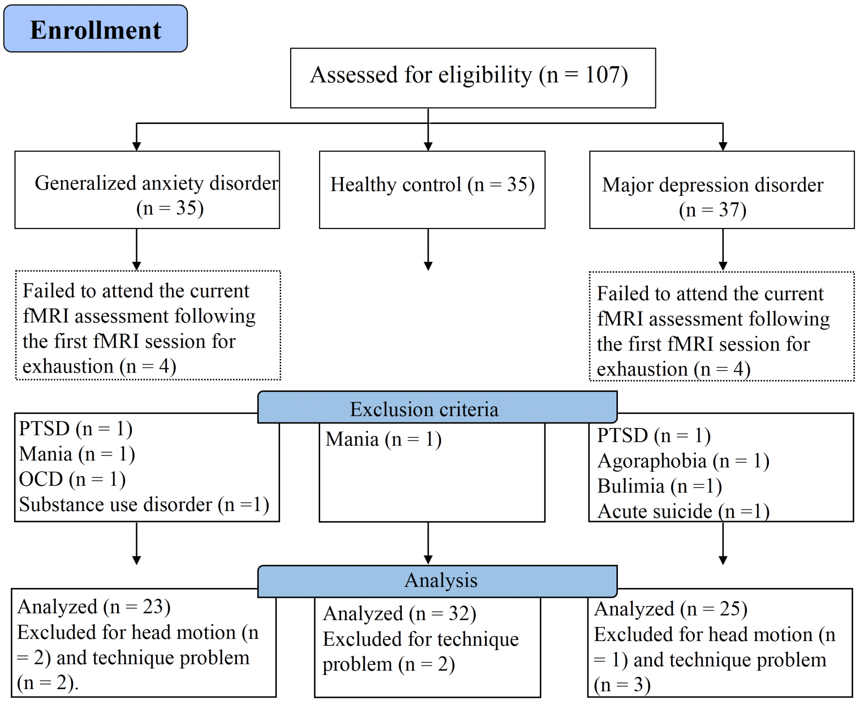
